# User-centred Design of a Clinical Decision Support System for Palliative Care: Insights from Healthcare Professionals

**DOI:** 10.1101/2022.06.03.22275904

**Authors:** Vicent Blanes-Selva, Sabina Asensio-Cuesta, Ascensión Doñate-Martínez, Felipe Pereira Mesquita, Juan M. García-Gómez

**Affiliations:** Biomedical Data Science Laboratory, Universitat Politècnica de València, Valencia, Spain; Polibienestar Research Institute, Universitat de Valencia, Valencia, Spain; Divisão de Hematologia. Departamento de Clínica Médica, Universidade Federal de Juiz de Fora, Juiz de Fora, Brazil

**Keywords:** CDSS, Design, Machine Learning, Palliative Care, User-centred validation, Usability

## Abstract

Clinical Decision Support Systems (CDSSs) could offer many benefits to clinical practice, but they present several adoption barriers regarding their acceptance and usability by professionals. Our objective in this study is to validate a Palliative Care CDSS, The Aleph, through a user-centred methodology, considering the predictions of the AI core, the usability, and the user experience. We performed two rounds of individual evaluation sessions with potential users. Each session included a model evaluation, a task test and a usability and user experience assessment. The Machine Learning predictive models outperformed the participants in the three predictive tasks. SUS reported 62.7± 14.1 and 65 ± 26.2 on a 100-point rating scale for both rounds, respectively, while UEQ-S scores were 1.42 and 1.5 on the –3 to 3 scale. Think-aloud methodology and the inclusion of the user-experience dimension allowed us to identify most of the workflow implementation issues.

## 1 Introduction

Clinical Decision Support System (CDSS) are computer systems designed to impact clinician decision-making about individual patients at the point in time that these decisions are made [1]. Interest and research about CDSS are motivated by their potential benefits documented in the scientific literature: increased patient safety by reducing medical errors or avoiding advice against protocol; improved service quality due to better adherence to guidelines, and increased service time dedicated directly to the patients; cost reduction by processing faster the demands and avoiding duplicated tests; improved administrative functions by incorporating elements such as automatic documentation; diagnosis support and workflow improvement [2, 3].

However, despite the multiple virtues that CDSSs could bring, there has been a lack of adoption of these systems into the clinical practice [4, 5, 6, 7, 8]. Several studies pointed to the main barriers to adoption, in which we could find two broad categories: socio-cultural factors and usability. Socio-cultural barriers refer to the beliefs of health care professionals or their organisation regarding the CDSSs, such as the idea of loss of autonomy, the feeling of being replaced by the system, low computer literacy, lack of trust in the system, failure to fulfil a perceived clinical need, legal uncertainties and misalignment between human needs and the technical system [9, 10, 11, 12]. Liberati *et al*.[10] proposed several strategies to deal with those barriers depending on the physicians’ beliefs regarding the CDSSs. Most of them are based on communication, training, and highlighting the system’s benefits.

On the other hand, usability barriers refer to the difficulties found by the user while using the software. The most common problems in this category are the difficulty to operate the software, the disruption of the workflow, the decrease of face-to-face time with the patients [2] and the alert fatigue due to excess notifications by the system [12, 13]. These challenges have been addressed previously by other authors by performing usability pilots with potential software end-users, mostly Healthcare Providers (HPs), in order to identify and correct the different usability problems of their CDSSs [13, 14, 15, 16].

Usability studies usually follow a general scheme. The participants are exposed to the software in a controlled environment and the session is usually taped and/or with the researchers taking field notes. Participants must try accomplishing tasks mimicking real scenarios, which in some studies receive the name on “near-live” simulations [17]. Think-Aloud methodology [18] is commonly used during the whole study, this method consists of asking the participants to express their doubts, opinions, and in general, any thought regarding their experience with the tool. Finally, the usability of the software is quantified through a scale or an index. One of the most popular evaluation tools is the System Usability Scale (SUS) [19, 20].

It is generally accepted that a positive User eXperience (UX) is essential to any software acceptance [21]. Despite existing a close relationship between usability and UX concepts, there are some differences worth studying, primarily related to the hedonic category [22], i.e., how ‘pleasurable’ the users find to use the software. In addition, UX also studies emotions, beliefs, preferences and perceptions. These concepts directly impact the adoption of a CDSS, concretely, they are strictly related to the previously mentioned socio-cultural barriers. Therefore, a UX study is essential to assess and improve technology adoption.

Another crucial aspect while maximizing the probability of a successful implementation of a CDSS in clinical practice is their design from the initial stages. An interdisciplinary team is highly recommended, including data scientists, programmers, usability and UX experts, the HPs as potential users of the software and other stakeholders such as representatives from hospital management to have a clear vision of the requirements [11, 23]. Planning a pleasant interface is also important since some studies reported users being more tolerant to minor usability issues if they found the interface visually appealing, which is known as the aesthetic-usability effect [24].

In our previous work [25], we developed a set of predictive models to assist the Palliative Care (PC) referral with hospital admission data on older patients using mortality and frailty predictions as main criteria. The result of that study was a demonstrator for a complete CDSS called The Aleph PC. Our study reported that these models accurately predicted which patients had a short survival time and were likely to become frail. Thus, our goal in this work is to validate the Aleph PC using user-centred techniques [26] and determine how different health professionals with PC experience envision the use of a PC CDSS in the clinical practice. First, we evaluated The Aleph PC’s mortality and frailty models against the HPs predictions to obtain a baseline, and second, we assess the usability and UX of the system alongside the different insights of the HPs on how to build an useful PC CDSS.

## 2 Materials and methods

### 2.1 The Aleph CDSS Platform

The Aleph PC is an open-access Machine Learning (ML)-based CDSS implemented as a web platform. The application is divided into three main screens; in the first one, the user introduces the different data required for the PC predictions, including administrative information, Barthel [27] and Charlson [28] indexes, laboratory results and a few diagnosis variables (Figure 1a). After completing the form, the results are calculated and displayed on another screen (Figure 1b); these results include a numerical result for each model and a Machine Learning (ML) explainability figure. We have used the Shapley Values (SHAP) [29] to display a graph with the relation between the input and the prediction obtained. Finally, the Files section (Figure 1c) allows the user to save the current case, load a different case or test the application with predefined test cases. The version tested in this study can be accessed here: https://demoiapc.upv.es/^1^.

**Figure 1:**
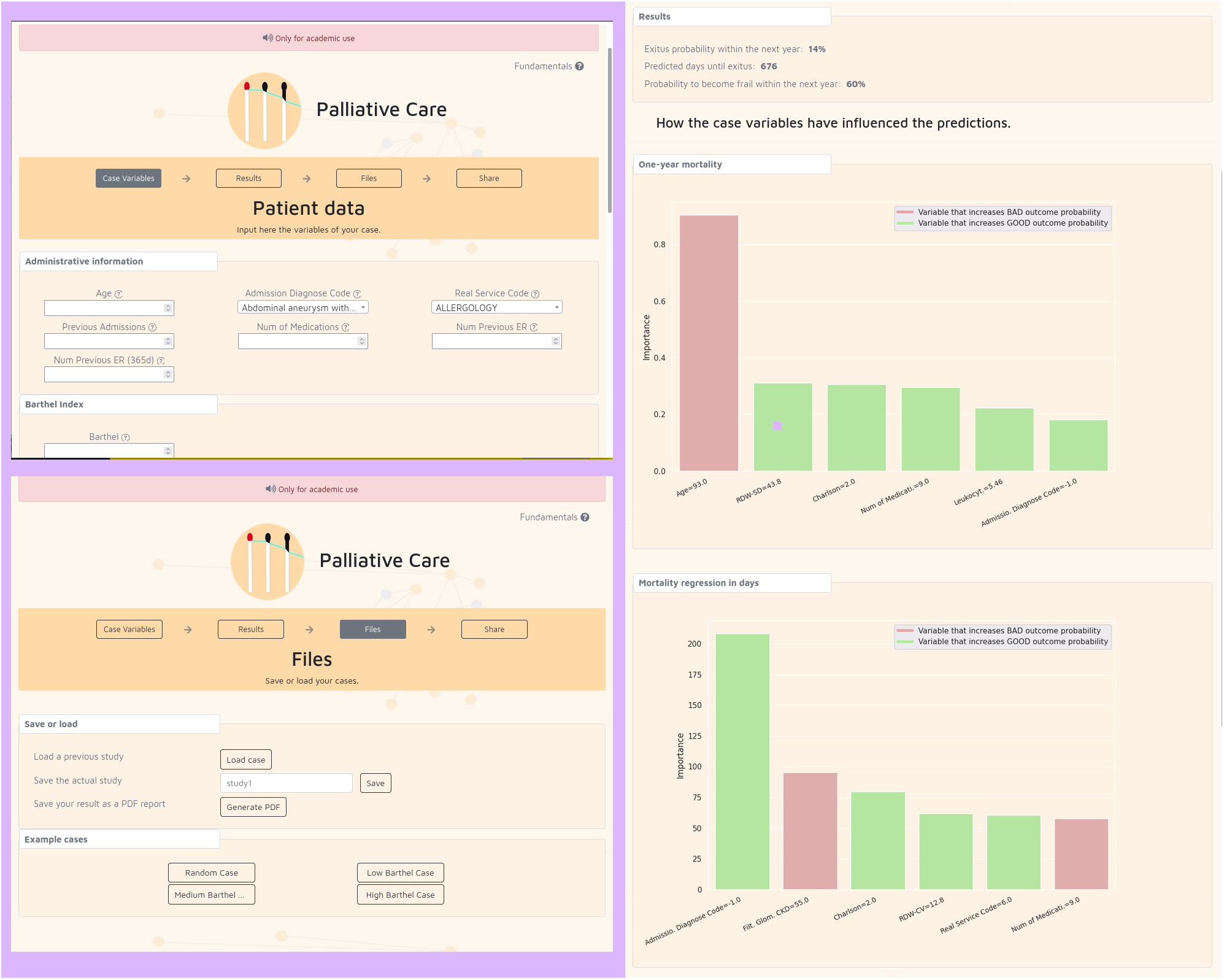
a) (left-top) Screen where the user inputs the data b) (right) Screen where the results are shown c) (left-bottom) Screen to manage the files and the predetermined examples

### 2.2 Recruitment process

We recruited healthcare professionals used to treat patients with bad prognostic within a wide variety of roles and possible end users of the CDSS. In concrete, we focused on: nurses, primary care physicians, hospitalist physicians, PC consultants and specialists like oncologists, neurologists or pulmonologists. This decision ensured the inclusion of different approaches working with complex patients in need of PC. The authors drafted a list of possible participants with no direct relationship with the development of The Aleph PC, and followed the snowball sampling technique [30]: once identified the first volunteers we asked them for other colleagues willing to participate until we completed our target sample size. Invitations to participate in the study were sent by email.

### 2.3 Study structure

#### 2.3.1 Participation

The study was defined as an iterative user-centred validation. Participants were invited to individual evaluation sessions where one of the team members acted as session guide. On some occasions, a second member of the team spectated the evaluation session and collaborated taking field notes. Evaluation sessions were performed by video-conference, where the participants shared their screens while interacting with The Aleph PC. The think-aloud methodology [17] was used during the whole session. The duration of each evaluation session was around one hour and their overall structure is displayed in Figure 2.

**Figure 2:**
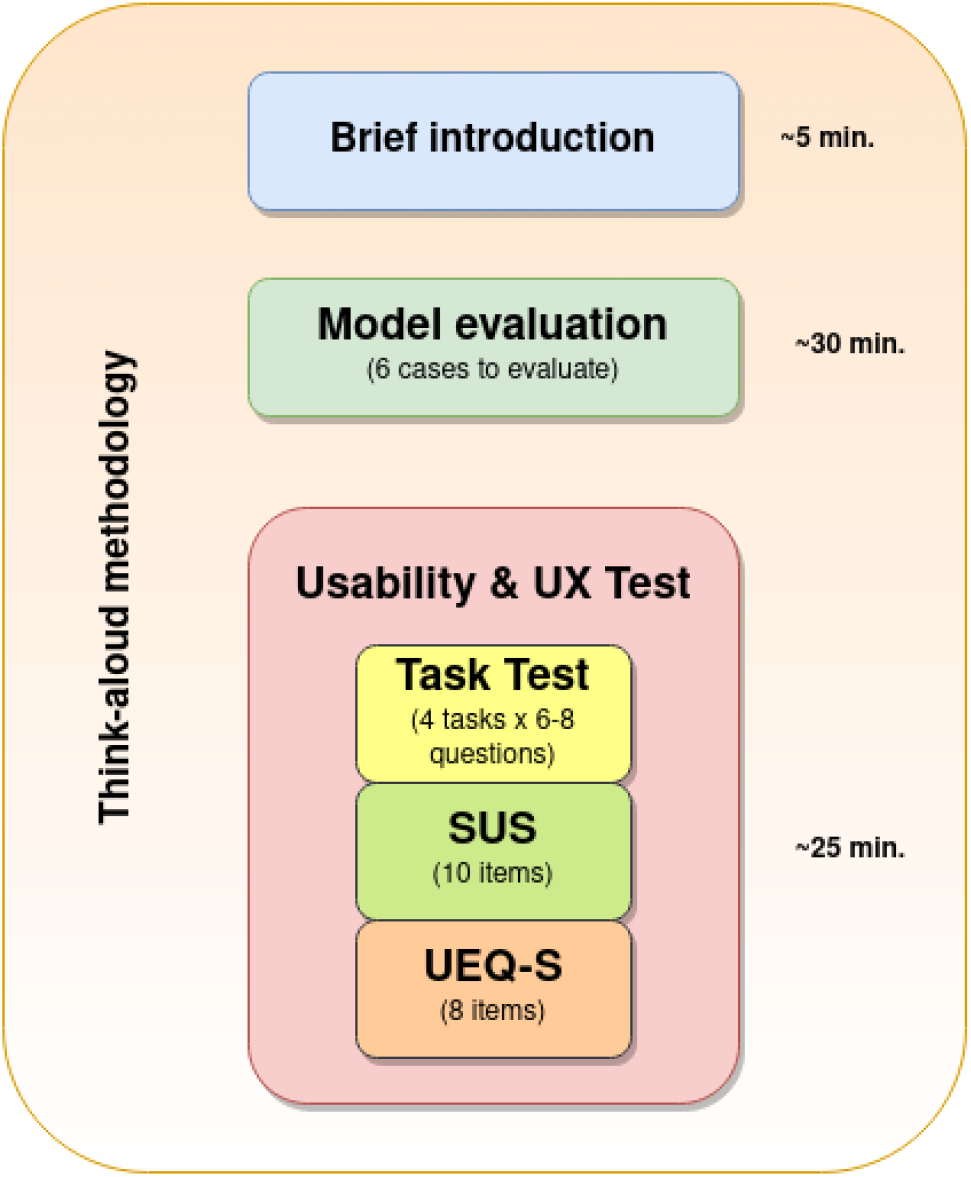
Overview structure of the sessions including three sections and the approximate time spent in each one of them: 1) Brief introduction, 2) Model evaluation and 3) Usability and UX test.

We defined two rounds of sessions, with a period of 15 days between them to adapt the software based on the feedback obtained during the first round. Sixteen participants were invited in the first round, 15 of them (93.75%) agreed to participate. For the second round, 8 participants were invited, and 6 finally responded (75%). We performed a greater number of sessions during the first round to detect as many usability problems as possible. We settled on six respondents during the second round due to the difficulty of finding participants and the fact that we already reached a number of participants that allow us to detect most of the usability problems according to the Nielsen Normal Group [31].

The distributions between the participants in both rounds was the following: fifteen of them were physicians with the following roles: 7 general practitioners, 5 hospitalists, 1 PC consultant, 1 oncologist and 1 neurologist. The other 6 participants were nurses. The distribution between sex was: 13 males (61.9%) and 8 females (38.1%). Distribution by country was Italy (5), Brazil (4), Spain (4), Greece (4), Scotland (2) and Portugal (2).

#### 2.3.2 Model validation

First, we implemented a model evaluation which was performed in an unlinked section^2^ of The Aleph PC. After introducing some basic information, the participants faced 6 vignettes: already filled, non-editable input forms containing real cases. Then, at the bottom of the page they were asked to fill in their own predictions regarding one-year mortality (yes/no), mortality regression (months interval) and one-year frailty. The same vignettes, in the same order, were asked for all 21 participants across both rounds. No time limit was established, and participants were asked to use any information resource (internet search, books…) they needed to make their own predictions.

We calculated the accuracy, sensitivity and specificity for the one-year mortality (1ym) and the one-year frailty (Frailty) models. Since we asked the participants for their predictions in months to facilitate their response, we had to transform the output of the regression model from days to months. Therefore, we divided the number of days by 30, discarding the decimal part, and then transformed it into an interval. The interval was determined to have a width of 4 months based on the results of our previous study, so we used the prediction in months ±2 months as interval bounds. We then calculated the accuracy of the participants and the model by checking if the real value of the cases in months belonged to the interval (lower bound ≤ real value ≤ higher value).

#### 2.3.3 Usability and UX validation

In the usability and UX section, the participants answered a Google Form questionnaire while they were testing the The Aleph PC. Participants were asked to do a ‘task test’: to perform four simple tasks using The Aleph PC and answer a series of questions after each of them. The tasks covered all the implemented functionality for the CDSS: 1) input a feasible case, 2) check the results and understand the graphics, 3) save the current case and, 4) load a case previously stored. The questions after each task interrogated the participants about the difficulty, the perception of time spent, the number of errors encountered by the participant (including unexpected behaviours and elements they did not understand) and the satisfaction obtained by performing the task. All questions were mandatory.

After the task test, we tested the usability and experience with the SUS questionnaire [19, 20] and the User Experience Questionnaire short version (UEQ-S) [32]. Both questionnaires were implemented into the same Google Form page. The participants were asked to stop sharing their screens during the completion of both tests.

## 3 Results

### 3.1 Model evaluation

The ML models outperformed the healthcare professionals’ predictions in both mortality and frailty (see Table 1). The mean width for the intervals provided by the participants in the regression prediction was: 16.2 months Confidence Interval (CI) 95% (13.5 to 18.9) against the fixed 4 months for the models.

**Table 1:**
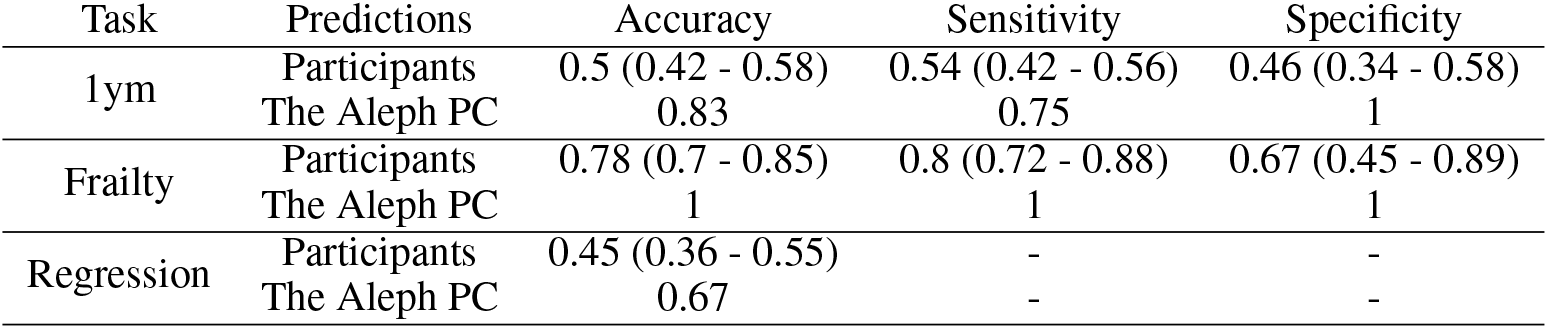
Summary of the metrics for the participants and the ML models in the three tasks for the 6 cases evaluated. Mean and 95% confidence intervals are reported per participant. ML are deterministic so no variability on the prediction was found.

### 3.2 Qualitative results

Regarding the qualitative results based on the think-aloud method and the authors’ notes on the participants’ behaviour, we created a list of improvements after each round, the changes were focused on interface details: removal of the Diagnosis Related Group variable because it could be inferred from the ICD9 code, replacement of the ICD9 codes by their name, improved tooltip descriptions, and added reference values for the laboratory variables. Table 2 in supplementary materials^3^ contains the complete log of changes introduced in both rounds. Most of the participants provided feedback regarding the subset of variables, suggesting other variables they are more familiar with or disregarding present variables as unimportant or unavailable in their workflow. The feeling towards the CDSS was primarily positive, and the idea of the PC identification using ML technology was well received. Few of the participants felt confused with the first interaction with the software but many of them affirmed they have learned to use it after the tasks test. Participants from hospital settings suggested picking automatically the diagnosis and laboratory results from the Electronic Health Records (EHR), whereas other participants did not care about complete integration due to the lack of system integration in their respective environments. We found a participant in each round who was sceptic about the use of computers for decision making and did not believe in the benefits of the technology, rating every aspect of the system very low and providing poor opinions about the system through the think-aloud method.

### 3.3 Performance of tasks

Figure 3 shows the distribution of the answers for both rounds. Almost every measured feature increased their percentage of positive feedback during the second round. The most significant improvement was on task four (load a case), where despite the increased number of errors, the perceived difficulty, time spent, and satisfaction improved.

**Figure 3:**
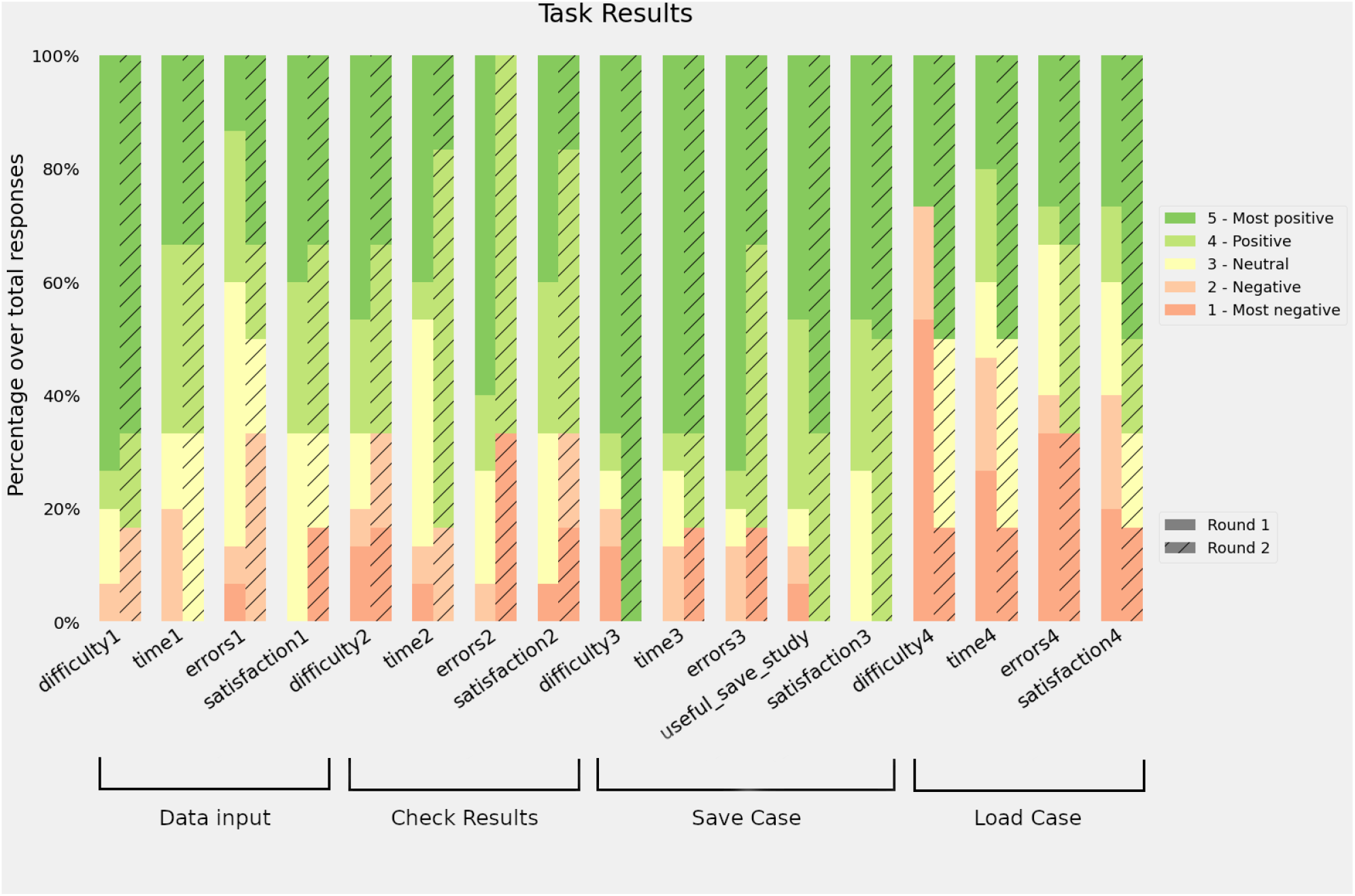
Results for the different tasks during the task test. Bars represent the distribution of the responses. A positive response means that the participant found the task: easy, short, with a low number of errors or satisfactory.

### 3.4 Usability

Responses to the 10 SUS items scores were recorded, all items were mandatory, so no missing values were present. The first round of the evaluation sessions obtained a mean of 62.7 ± 14.1, while the second round increased its score to 65 ± 26.2. The distribution of the answers for the different items can be found in Figure 4. We have used the adjusted scores instead of the raw scores for all items to help with visualization. Round 2 has a greater proportion of positive responses in 6 out of 10 items, nonetheless the first round has a lower score and lower standard deviation.

**Figure 4:**
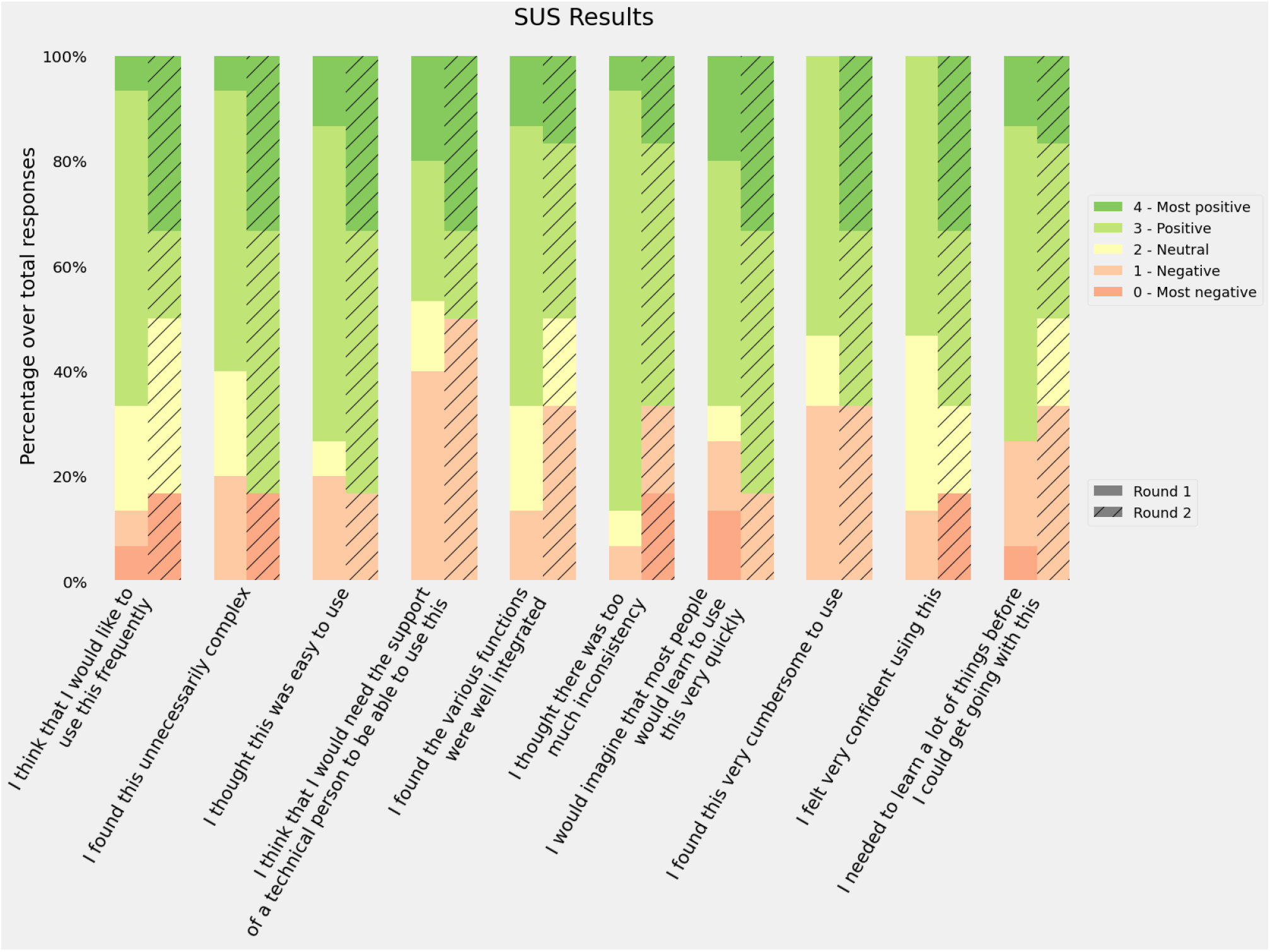
Responses to the SUS questionnaire. Bars represent the distribution of the responses using the adjusted scores: raw scores minus 1 for items in the odd position; 5 minus raw scores for items in an even position.

Previous studies have tried to map intervals of the score into categories such as “Poor”, “OK” or “Good” or school grading scales [33] in order to provide a better usability reference. According to these frameworks, our results for both rounds would be classified as D (lowest passing score), the first round as “OK - low marginal acceptance” and the second round as “OK - high marginal acceptance”. However, if we recalculate the SUS average score excluding the sceptical participants, the average rating would be 63.9 ± 13.8 and 74.5 ± 16.8 which are D “OK - High marginal acceptance” and C “Good - Acceptable”.

### 3.5 User experience

Answers to the UEQ-S questionnaire were recorded, with all items being mandatory. Figure 5 shows the distribution of the responses for each item in the questionnaire. The median for the second round was always greater than the first round, and the average scores were: 1.4 in the first round and 1.5 in the second one. The Pragmatic score was slightly higher in the first round (1.3 vs 1.2) and the hedonic score improved during the second round (1.5 vs 1.8).

**Figure 5:**
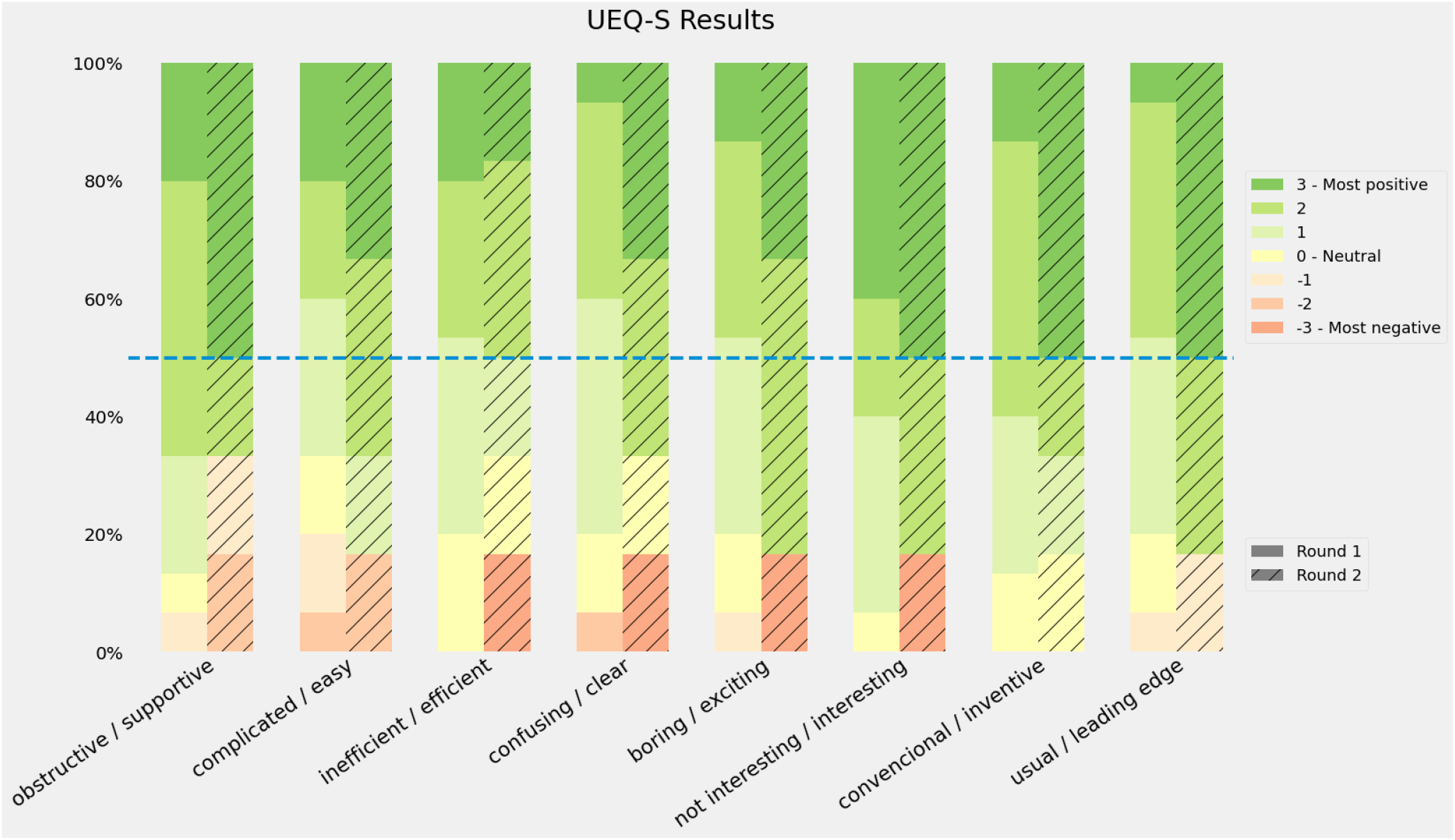
Results for the UEQ questionnaire. Bars represent the distribution of the response. Positive responses mean that the participants agreed with the positive quality of the software (supportive, easy…)

Authors of the UEQ-S provide a benchmark in order to compare the study results, despite this benchmark being intended for the full-size UEQ, the results may be acceptable to estimate how good the user experience is. Figure 6 shows the results of both rounds in the three categories and their benchmark score.

**Figure 6:**
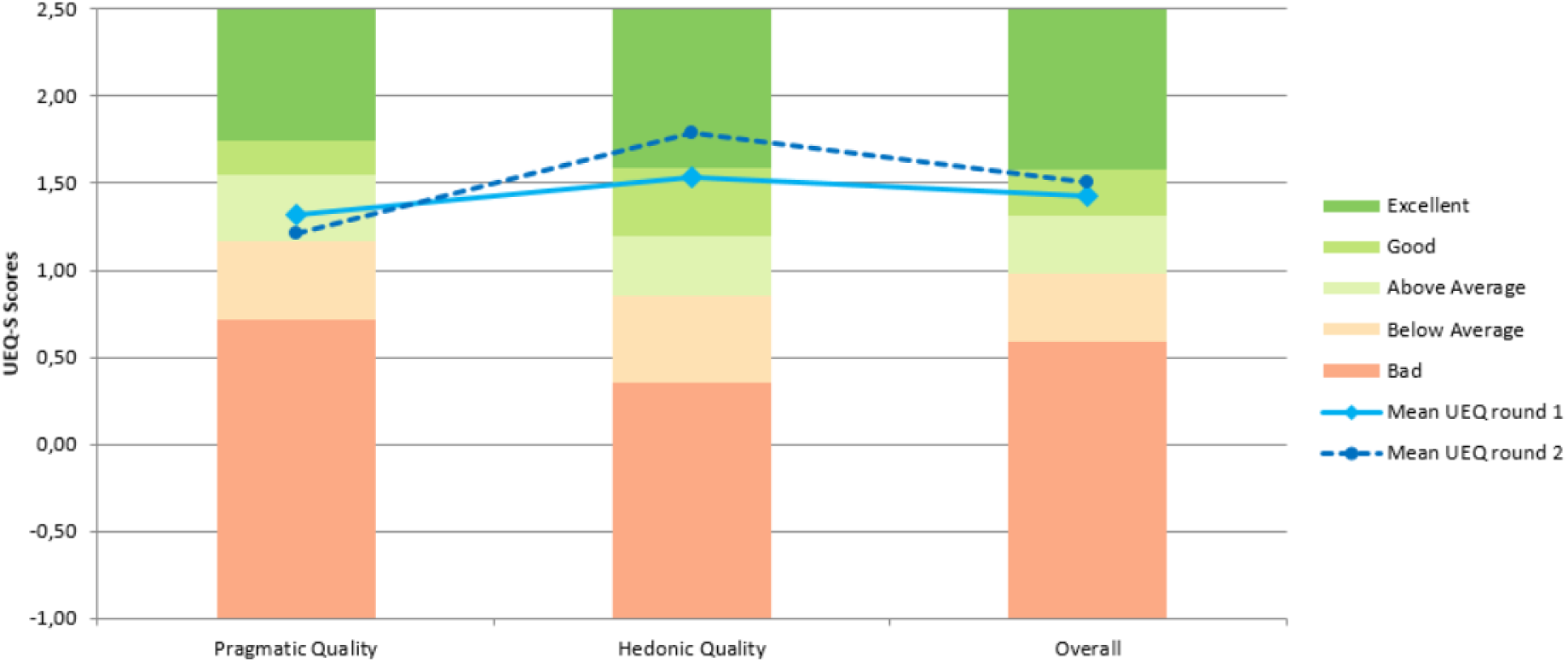
UEQ results by categories against the official benchmark. The grade assigned to each category depends on the results from different studies used to create the benchmark.

## 4 Discussion

In this study, we performed an iterative user-centred validation of a CDSS aimed to support healthcare professionals in the identification of patients in need of palliative care. This two-round validation process involved decision, usability and user experience tests. During this study, the predictions provided by the models were more accurate in both sensitivity and specificity metrics for both classification models than those provided by the participants. The regression model accuracy result depends on the width of the interval; we selected 4 months as an acceptable error based on the original reported mean absolute error [25]. Other studies have described the low accuracy of clinicians when predicting one-year mortality through mechanisms like the Surprise Question [34]. However, our intention with this comparison was to set a reasonable baseline for the predictive models. There are several factors that play against the performance of clinicians in this evaluation 1) they are not good at taking decisions over EHR data [35], 2) not having physical access to the patients affect HP’s intuition [36] and 3) the number of cases to evaluate was low. However, these results indicate that The Aleph PC could help improve the clinicians’ predictions with data.

The results of the task test indicate that the four tasks were not perceived as difficult. Task 4 (load a file in the platform) presented the highest number of negative responses regarding its difficulty during the first round, but it improved at the second round after we relocated the load button from the input variables section to the files section, as suggested by the participants. The tasks were also perceived as fast to carry on, despite the need to input all the variables manually in the first task. Levels of satisfaction were high, increasing in the last two tasks for the second round after the interface improvement. It is worth mentioning that all participants in the second round rated positively the option to store the data into a csv file so they would be able to revisit the case later.

The different SUS dimensions were considered positive (scores 3 and 4) by at least half of the participants in both rounds. The best scores were related to the perceived difficulty: “I found the system unnecessarily complex”, “I thought the system was easy to use” and “I would imagine that most people would learn to use this system very quickly”. Consistency was also rated among the best (“I thought there was too much inconsistency in this system”). The worst scores were related to the confidence of the participants (“I found the system very cumbersome to use”, “I felt very confident using the system” and “I think that I would need the support of a technical person to be able to use this system”). These results suggest that participants found The Aleph PC easy to use but containing some elements that they were not familiar with and/or needed further explanation. The average score was “passing” though there is room for improvement according to participants’ feedback.

UEQ-S results reported in Figure 5 provide two main interpretations, first, the median UX scores were higher in the second round after the introduction of improvements. Second, most of the dimensions kept their values on neutral (0) and positive (1, 2, 3) values. Some outliers responses can be observed especially on the second round corresponding with the sceptical participant, which means that their experience with PC CDSS was positive, a deep look into the supportive category is needed to address possible undetected issues.

After analysing the results from SUS, the UEQ-S and the comments obtained from the think-aloud methodology we obtained a rich vision of the perception of the application. It seems that most of the participants were predisposed to use a tool such as The Aleph PC to obtain prognosis predictions that could influence PC action. Even the two participants who expressed their dislike of the CDSS and showed scepticism about the tests had their own vision about how the application should be: whereas the first-round participant did not feel that the tool was useful for their service, the second-round participant specified *“The tool is boring. The end-of-life idea should be obtained in 10-15 seconds”*. Only a few participants had experience using CDSSs, but all of them understood these kinds of applications as supporting tools instead of as a threat to their autonomy [10, 37, 38]. In addition, all of them understood the difficulty in identifying early PC patients so they were prone to accept a tool that may help them.

Although, the usability results were not as positive as the hedonic quality. Some hypotheses could be extracted from the sessions. Mainly, The Aleph PC did not show a perfect fit in its current status to the different participant backgrounds. Participants from non-hospital environments commented on how some of the variables required by the models did not match with the information managed by their centres. Being unfamiliar with some required input could have a detriment on the perceived ease of use of the product. Four of the participants commented on the nature of the data introduced, they considered that introducing historical data in the application instead of a “snapshot” of a given moment could be better for the prediction. Also, a couple of the hospital physicians made suggestions about automatising the input and the output and their integration with the EHR following the schema of integrative CDSS [11]. These findings are in line with the concept of unremarkable AI [35], where the AI systems are meant to be integrated in the current workflow and did not disturb or overwork the HPs.

As stated by other works on validation, usability is a key factor of the success of the CDSS implementation. Usability tests based on the performance of tasks are sometimes described as “near-live” simulations, and the posterior usability assessment is the standard to discover usability issues and improve the final product [13, 39]. Nonetheless, developing a perfectly usable application does not guarantee its implementation success since there are a list of socio-cultural barriers in the adoption of these technologies [10] which are directly related to the vision and opinions of the physicians and their organisations regarding these products.

The inclusion of the UX test during the evaluation sessions allowed to detect participants’ predisposition and feeling towards The Aleph and the general idea of using a CDSS as a daily tool in clinical practice. Despite the diversity of backgrounds among the participants, there was an agreement on the usefulness of the tool. Also, the whole set of participants, with two exceptions, believed in the technology and the evidence behind the predictive models. This is especially relevant since trust has been detected as one of the main issues regarding the CDSS acceptance [10].

The inclusion of two rounds allowed us to test if the changes implemented after the first set of sessions influenced the usability and UX during the second round. The difference between SUS and UEQ-S overall scores were not significant using the T-Test (P>0.05). However, we observed an improvement in certain dimensions of the metrics. We could not extract valid comparisons per role due to sample size restrictions since most of the nurses were in the second round, and most of the physicians were in the first round. At the same time, those groups were not homogeneous and contained HPs working in hospitals, primary care centres, external services and rural environments. Regarding the number of iterations, we could have set a bigger number in order to ensure that the minimum number of issues is kept in the software. However, the changes identified during the second round were either detail such as the use of abbreviations and acronyms or barriers derived from the data source from which the models were created. Therefore, we considered that most of the fixable issues were identified and we didn’t need to perform any extra iterations.

The participation of professionals with different roles and backgrounds allowed us to observe the diverse needs in the highly heterogeneous PC implementation and workflows. There are significant differences between inpatient and outpatient settings [40], medicine specialities [41, 42, 43], and urban or rural environments [44]. In those different contexts, physicians and nurses play different roles in the PC needs identification. For example, in the work by Zemplényi *et al*.[45] the authors describe how the nurses are the first to detect the needs and then the cases are discussed with the physicians. This may be different in other settings, such as rural areas where physicians visit older patients. In our study, we observed that participants working in non-hospital environments were more concerned about the availability of the variables, especially the laboratory results. Both physicians and nurses thought they could benefit from The Aleph in their workflow.

Another relevant detail in our implementation is the inclusion of ML explainability in the system (Figure 1b). ML explainability could defined as the human quality to understand the relation between the system input and their predictions [46] and has been proposed many times as a solution to one of the most common CDSS adoption barriers, as stated by Shortliffe and Sepúlveda “black boxes are unacceptable” [47]. CDSSs should be transparent to the user to allow them to accept or dismiss the prediction or recommendation. However, recent studies have highlighted possible problems when trying to create explainability mechanisms to single predictions. In its viewpoint, Ghassemi *et al*.[48] discourage their implementation as patient-level systems. Since we received positive feedback from the participants on this feature, we decided to not remove the explainability graphs after the second round. However, we acknowledge the need for further study for these kinds of features and their impact on the clinical workflow.

Since the start of this work, our team has followed the design recommendations from previous studies, focusing on two main aspects: team composition and interface design. Our team included multiple roles: physicians, designers, usability and UX experts, ML researchers and programmers. This is especially relevant since a multidisciplinary team can get a better understanding of the real requisites of the project and mitigate workflow disruption [11, 23]. The interface was carefully designed, the layout was implemented focusing on usability and the colours used were extracted from the PC logo that was created previously by an artist. As described in [24], the aesthetic part of the application has a direct effect on the perceived usability, therefore an effort to create a visually attractive application must be set in place. The scores obtained in the UEQ-S hedonic category reflect the acceptability of the aesthetics, however, none of the participants commented explicitly on the visual aspect.

The main strength of out work was that our methodology assisted us to obtain the insight of different pitfalls identified in previous works [2, 9, 10, 11, 12, 13] using HPs’ insights. Through the usability test we discovered that the system is *good enough* for the participants. However, concrete changes are needed depending on the context where the CDSS is deployed to maximise the usability aspect. With the predictive models being evaluated in a previous publication [25] and the evaluation of usability and UX in this work we followed an exhaustive user-centred validation path. We created anecdotal evidence to support the UX dimension within the standard usability tests. In addition to this, we managed to get a very diverse sample of participants in terms of roles and countries, providing us with a richer version of the health providers’ needs regarding PC in different countries.

Our work also presents some limitations. First, the model evaluation was performed with a low number of participants, and since the ML models are deterministic once trained, the evaluation on the machine side is only on the six different cases. However, with respect to the accuracy of the predictions, this is not a problem since the models were evaluated previously. Another limitation in our study is the requirement of manual input of hospital admission data because it is disconnected from the EHR. This could present a perk in roles such as primary care physicians working in rural areas but breaks the premise of automatizing the data collection as much as possible and increases the possibility of human errors. In addition, in this demo, we have not addressed some problems regarding the variability, temporal and related to different medical centres, over data distributions [49].

As future work, we would like to adapt the tool to the different roles and clinical workflows we have identified. A further study focused on the different PC roles and their needs regarding The Aleph PC would be needed to provide a perfect fit and improve usability. Further adaptations and validations of the ML models would be needed to ensure the models keep their predictive power in other populations. In order to go further with The Aleph PC, we would need to create a pilot for potential users to incorporate the tools in their daily routine and gather long-term feedback. Further research about the ML explainability and reportability in needed to create a transparent and auditable system that could improve the acceptance of the technology by helping avoid legal problems [11, 46]. In addition, a study focused on the mortality and frailty prediction accuracy by HPs may be needed to estimate a fair baseline for predictive models to improve.

## 5 Conclusions

Our main findings indicate that the predictive models have performed better than the baseline composed of the HPs predictions. The system presents great UX hedonic qualities, i.e., participants were excited to use the tool, and they rated positively the fact of having helped to identify patients with bad prognosis. They did not feel their independence threatened by the Aleph PC. Performance regarding usability was modest but acceptable. Based on the notes taken during the think-aloud methodology, the authors hypothesise that the usability scores for the current version are maximised and would only improve if the tool was adapted for the different roles and contexts represented in the participant’s sample. We have created anecdotal evidence that an iterative user-centred validation, including UX, provides a broader vision to address CDSS acceptance issues. The objective of The Aleph PC is to step further in the objective PC criteria inclusion.

## Supporting information

Suplementary table including the changelog for the platform

## Data Availability

The data that support the findings of this study are available from the corresponding author, V.B.S, upon reasonable request.

## 6 Declarations

### 6.1 Ethical Approval and Consent to participate

This study was assessed and approved by the Ethical Committee of the University and Polytechnic La Fe Hospital of Valencia (registration number: 2019-88-1).

### 6.5 Authors’ contributions

V.B.S, S.A.C and A.D.M designed the evaluation sessions. V.B.S, S.A.C and F. P. M performed the evaluation interviews. V.B.S, A.D.M and J.M.G.G drafted the manuscript. S.A.C, A.D.M, F.P.M and J.M.G.G. performed a critical review of the manuscript.

## 6.6 Acknowledgements

The authors thank their help during the recruitment to Dr. Michael Doumas from and Henrique Silva. Special thanks to Dr Gordon Linklater for his contributions.

## Footnotes

1 Last accessed June 29, 2022

2 http://demoiapc.upv.es/validations/launch_validation Last accessed June 29, 2022

3 https://www.medrxiv.org/content/medrxiv/early/2022/06/05/2022.06.03.22275904/DC1/embed/media-1.xlsx?download=true

